# Distribution of rare *LOXL1* missense alleles, haplotypes, and diplotypes suggests association with reduced risk of glaucoma-related exfoliation syndrome

**DOI:** 10.1101/2021.08.05.21261676

**Authors:** Inas F. Aboobakar, Rob P. Igo, Tyler G. Kinzy, Jessica N. Cooke Bailey, Chiea Chuen Khor, Tin Aung, Robert Ritch, Arthur J. Sit, Richard K. Lee, Margaret Pericak-Vance, Jae H. Kang, Jonathan L. Haines, Louis R. Pasquale, Janey L. Wiggs

## Abstract

**Purpose:** Common *LOXL1* protein-altering variants are significant genetic risk factors for exfoliation syndrome (XFS) and the related secondary glaucoma (XFG). A rare *LOXL1* missense allele was associated with protective effects in a Japanese cohort, suggesting that other rare alleles may also exhibit protective effects. The goal of this study is to assess the contributions of rare *LOXL1* variants to XFS/XFG risk in cases and controls from the United States (US).

**Methods:** *LOXL1* rare variants (minor allele frequency (MAF) < 1%) were identified from Human exome BeadArray (Illumina) data for 1118 XFS/XFG cases and 3661 controls (Mass Eye and Ear (MEE) cohort) and from exome sequence data for 284 cases and 37,499 controls in *All of Us* (AoU) The distribution of rare variants, haplotypes, and diplotypes was examined using Fisher exact test.

**Results:** Four rare *LOXL1* missense alleles, more common in controls, were identified in MEE, P= 7.6E-4), and 456 variants identified in AoU were found only in controls (P=0.045). The rare protective alleles were preferentially located (P= 5.8E-45) in a LOXL1 intrinsic disordered region (IDR) potentially involved in LOXL1 aggregation. Haplotypes that included the rare or minor variants were more common in controls compared to cases in both MEE (Odds Ratio (OR)= 0.21 (95% Confidence interval (CI): 0.19-0.24), P=1.7E-173) and AoU (OR=0.28 (95% CI: 0.23-0.34), P=4.4E-41), and heterozygous diplotypes were also significantly associated with reduced risk overall in both MEE (OR= 0.45 (95% CI: 0.52-0.39), P= 1.7E-89) and AoU (OR=0.26 (95% CI: 0.18-0.33, P=7.6E-28). Diplotypes comprised of only homozygous genotypes were associated with increased disease risk in both MEE and AoU (OR= 4.16 (95% CI: 3.60-4.76), P= 4.2E-89 and 5.26 (95% CI: 4.12-6.73), P=1.87E-42, respectively) and excess homozygosity and decreased heterozygosity was correlated with disease risk for common *LOXL1* variants across multi-ethnic populations (P=1.0E-6).

**Conclusions:** Using exome array and exome sequence data from XFS/XFG cases and controls from the United States, we identified rare protective *LOXL1* missense variants and show that the distribution of the corresponding haplotypes and diplotypes was associated with lower risk of XFS/XFG. Diplotype results also demonstrated that *LOXL1* allelic heterozygosity was inversely associated, while homozygosity was associated with increased disease risk. These results suggest that *LOXL1* MAF variation among populations, with corresponding variation in genotype heterozygosity and homozygosity, determines the XFS/XFG association.

## INTRODUCTION

Exfoliation syndrome (XFS) is a systemic disorder characterized by progressive accumulation of abnormal fibrillar protein aggregates that can obstruct drainage of fluid from the eye, raising intraocular pressure (IOP) and causing exfoliation-related secondary glaucoma (XFG). XFG is the most common secondary glaucoma worldwide and is a major cause of irreversible blindness (Nazarali, 2018). XFS is also associated with premature cataract formation and complications during cataract surgery. Systemic conditions have also been associated with XFS, including cardiovascular disease (Chung, 2018), sleep apnea (Shumway, 2021), and obstructive pulmonary disease (Taylor, 2019).

*LOXL1* (lysyl oxidase-like 1) is a major genetic risk factor for XFS/XFG in all populations examined (Aung, 2017), with *LOXL1* risk variants occurring in up to 98% of patients (Fan, 2011). Aggregated LOXL1 protein is one component of the exfoliation fibrillar material (Zenkel, 2014; Sharma, 2009) that has been identified in extracellular spaces throughout the body, but in particular in the ocular anterior segment (Schlötzer-Schrehardt, 2009). The LOXL1 protein includes several intrinsic disordered regions (IDRs) that could be sites of initiation of protein aggregates, and recent studies suggest that deletion of the most disordered regions reduces protein aggregation (Bernstein, 2019).

Two of the major *LOXL1* risk variants, (G153D and R141L) are common missense alleles located within the IDR with a high likelihood of aggregation (Bernstein, 2019). (**Figure 1**). Of interest, in most populations studied, the common allele ‘G’ at 153 and the common ‘R’ allele at 141 are associated with increased risk; however, in East Asian populations the ‘L’ allele at 141 is associated with increased risk (Hayashi 2008), and in South Africans, the ‘D’ allele at 153 is associated with increased risk (Williams 2010). The R141 ‘L’ and G153 ‘D’ are the ‘common’ alleles in East Asian and South African populations, respectively. Several studies have investigated the role of *LOXL1* genetic variants including effects on protein function (Sharma 2016), gene expression (Pasutto, 2017), and dysregulation related to a long noncoding RNA also located within the LOXL1 genomic region (Hauser 2015). However, none of these studies have observed a consistent effect for the associated risk allele in all populations, which limits the determination of the underlying disease-causing mechanism(s).

**Figure 1.**
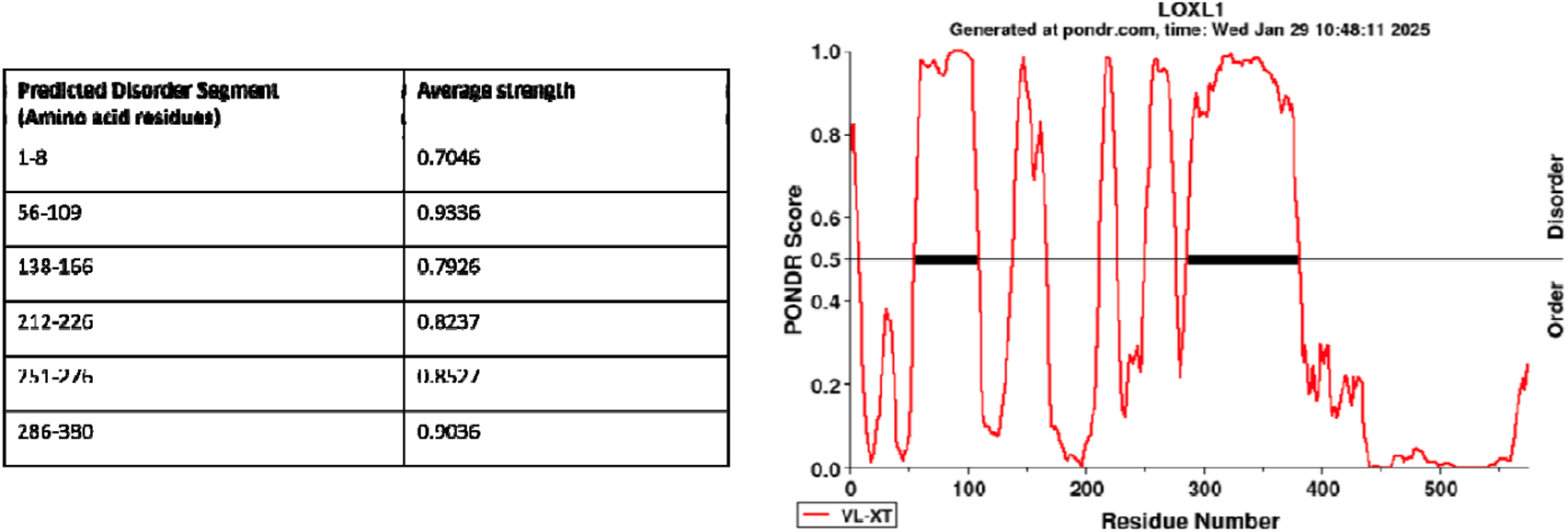
LOXL1 intrinsic disordered domains. Six disordered domains are predicted by the program PONDR (Predictor of Natural Disordered Regions) (Peng, 2005). Disordered probably greater than 50% is considered to be significant. The regions resulting in reduced aggregation when deleted were residues 28-95 and residues 125-185, Bernstein et al. (Adv Protein Chem Struct Biol. 2019;118:273-288).

A rare *LOXL1* protein-altering variant has been associated with significantly reduced risk of XFS in Japanese cases and controls (Aung, 2017). This finding, as well as the observation that the less frequent allele at G153D and R141L is protective in all populations studied so far (i.e., the heterozygous genotype is consistently associated with reduced risk), suggests that other rare or low-frequency alleles may also be protective. To investigate this hypothesis, we examined the effects on risk of rare *LOXL1* coding variants in two cohorts from the United States: a cohort of XFS cases and controls with European genetic ancestry from Mass Eye and Ear (MEE), as well as a cohort of XFS cases and controls with European genetic ancestry from the *All of Us* (AoU) Study.

## METHODS

### Massachusetts Eye and Ear cohort

This study adhered to the tenets of the Declaration of Helsinki and has been reviewed and approved by the Institutional Review Boards of the Massachusetts Eye and Ear, Harvard School of Public Health, and the Brigham and Women’s Hospital. Informed consent was obtained from the participants after explanation of the nature and possible consequences of the study. XFS/XFG cases were recruited from MEE (Harvard), Mayo Clinic, Bascom Palmer Eye Institute (University of Miami), the Nurses’ Health Study (NHS), and Health Professionals Follow-up Study (HPFS). The controls were previously genotyped samples from the NHS and HPFS, and were over the age of 40, self-reported Caucasians with European ancestry, and located throughout the United States (Kang 2016). Controls have no evidence of XFS by clinical exam or medical record or have repeatedly denied a self-report of glaucoma on biennial questionnaires administered over two decades. We also accessed genotype data from the MEE and NEIGHBOR controls (Wiggs, 2012; Margeta 2020).

All XFS cases were of self-reported European ancestry and were over the age of 40 and have documentation of characteristic ocular exfoliation material at the pupil margin or surface of the ocular lens, either through clinical exam or medical records. Clinical examination included measurement of visual acuity and intraocular pressure, slit lamp biomicroscopy, and fundoscopy. Cases also had visual field assessment primarily using the Humphrey automated visual fields. Individuals were excluded if other types of glaucoma (pigment dispersion, steroid-induced, uveitis) were evident on exam. All cases provided blood samples for DNA extraction.

### Genotyping and quality control

Cases were genotyped at the Center for Inherited Disease Research (CIDR; http://www.cidr.jhmi.edu/) using the Illumina OmniExpress+Exome platform that includes 700,000 common SNPs (MAF >0.3) and 250,000 rare or low frequency functional exonic SNPs collected from 12,000 exomes (http://genome.sph.umich.edu/wiki/Exome_Chip). The XFS/XFG controls were previously genotyped using the Omni Express+Exome platform. The NEIGHBOR control data set was also genotyped using the Illumina HumanExome BeadArray (Illumina, Inc., San Diego, CA) at CIDR.

Quality control for human exome array genotype data (Igo 2016) for all cases and controls was carried out as follows. The Illumina Genome Studio (Illumina) and PLINK (Purcell 2007) were used for all quality controls (QC) steps except where noted. Basic QC for samples included screens for call rate (≥98.5%) and high (≥95%) concordance with a previous Illumina 660K panel run on the same sample, where available (about 80% of samples). We verified recorded sex in the clinical records with genotyped sex by two criteria: mean fluorescence intensity on the X and Y chromosomes, plus genotype heterozygosity on the X chromosome and call rate on the Y, allowing male and female samples to have heterozygous X-linked and successful Y-linked genotypes, respectively. We tested samples for pairwise relationships and unexpected duplication using KING (Manichaikul, 2010).

We verified European ancestry from the first two principal components derived from genotypes at 9000 ancestry-informative markers by means of the SNP weights program (Chen, 2013), including representative HapMap CEU, YRI, CHB, and JPT samples as reference populations. Moreover, we conducted a principal components analysis over 52,040 independent (pairwise *r*^*2*^ < 0.1), common (MAF ≥0.005) SNPs using the smartpca program in EIGENSOFT (Price, 2006) to detect finer population structure.

Initial QC screens for markers included call rate (≥98%) and consistency with Hardy-Weinberg proportions (*P* > 10^−6^ by Fisher’s exact test). We screened markers for differences in allele frequency between whole-genome-amplified DNA samples and all other samples by the Fisher’s exact test and removed from analysis all markers with *P* < 0.0001. All pseudoautosomal, Y-linked, and mitochondrial SNPs were subject to review in Illumina Genome Studio (Illumina, Inc.), and if necessary, were manually re-clustered. We also confirmed genotype clustering for rare (MAF <0.02) variants from fluorescence intensity data by means of zCall, run with a stringent *z* score threshold of *z* = 21 for calling heterozygous genotypes. Every rare SNP with two or more additional heterozygous calls by zCall than by GenCall was reviewed in Genome Studio, and if necessary, cluster locations were adjusted manually. Data for four rare coding variants (G120S, S159A, A160P, S427F) included on the genotyping platform were extracted for analyses.

#### All of Us

Informed consent was obtained for all participants enrolled in AllofUs. The AllofUs IRB follows the regulations and guidance of the National Institutes of Health Office for Human Research Protections for all studies, ensuring that the rights and welfare of research participants are overseen and protected uniformly (AllofUs Research Program, 2019; AllofUs Research Program, 2024). Cases were older than 55 years and had an ICD9/10 code for XFS or XFG (366.11, 365.52, H40.14, H40.143, H40.149, H40.141, H40.142, H40.1490, H40.1430, H40.1431, H40.1432, H40.1413, H40.1411, H40.1433, H40.1421, H40.1434, H40.1422, H40.1491, H40.1423, H40.1412, H40.1494, H40.1492, H40.1420, H40.1493, H40.1410, H40.1424, H40.1414). Controls were older than 75 years and had no ICD-9/-10 code for any form of glaucoma, glaucoma suspect, or ocular hypertension. After data quality control (relatedness, sex mismatch), 371 XFS/XFG cases and 37,522 controls were identified. Since the majority of the cases were of European genetic ancestry, other genetic ancestries were removed, leaving 284 cases and 37,499 controls.

1190 rare variants (Minor allele frequency (MAF) <1%) identified in exome sequence data from the region corresponding to *LOXL1* coding sequence (Chr15, 73926784-73951837) were filtered to remove variants with allele counts of <2 in cases and controls and missingness >2%, leaving 47 variants which were mainly missense alleles and all only present in controls and with 456 controls carrying at least one rare allele.

### Other genetic ancestry-defined groups

For replication purposes, data from genetic ancestry-defined groups where the variant of interest was identified in Aung et al. (Aung, 2017), was included.

### UK Biobank

While diagnostic codes for XFS or XFG are not included in the UK Biobank clinical showcase, and therefore cannot be used for case or phenotype selection, the individuals with overall glaucoma codes (H40) would include these individuals. Using publicly available data (Genebass), we investigated overall burden results for the glaucoma groups as well as single variant associations.

### Multi-ethnic ancestry groups from literature review

To investigate deviations from Hardy-Weinberg equilibrium for the common risk alleles (G153D and R141L) we used study-specific data for 40 independent studies published as a meta-analysis by Li et al., (2021), that included European Caucasians, Asians, and Africans ancestry groups.

### Analyses

Distribution of the four rare *LOXL1* variants in MEE cases compared to controls and the 456 AoU variants in cases and controls was assessed using Fisher’s exact test. Haplotypes were constructed using IMPUTE2 (Howie 2009), and haplotype distribution between cases and controls was also assessed using Fisher’s exact test for both cohorts. Diplotypes were identified using the haplotype data for each study participant. Chi-square and Fisher’s exact test were used to assess Hardy Weinberg disequilibrium and variation percent homozygosity and heterozygosity with disease risk for common *LOXL1* risk alleles. To assess Hardy Weinberg equilibrium for the common risk alleles (G153D and R141L), genotype results from individual studies included in a large meta-analysis (Li, 2021) were tallied for each genetic ancestry group (Europeans, Asians and Africans). Using the frequency for the common and minor allele for each variant, the expected Hardy Weinberg distribution was calculated using p2 +2pq + q2 =1. The observed genotype distribution was compared to the expected using the chi-square test.

## RESULTS

For the MEE cohort, after sample and genotyping quality control, genotype data was available for 1,118 cases and 3,661 XFS/XFG controls. Additionally, genotype data was available for 2,606 primary open angle glaucoma (POAG) subjects without XFS/XFG from the NEIGHBOR study (Wiggs 2013). Cases and controls have similar proportions of females (**Supplementary Table 1**). The cases are on average older than both control cohorts; however, the overall difference was not significant (P= 0.26). Similar age and sex distribution was observed for the AoU cases (N=284) and controls (N=37499) (**Supplementary Table 1**).

### Distribution of rare missense alleles in XFS cases and controls suggest protective effects

In the MEE cohort we identified four *LOXL1* missense alleles with minor allele frequencies (MAFs) less than 1%: G120S, S159A, A160P and S427F (**Table 1**). Three of these have Combined Annotation Dependent Depletion (CADD) (Kircher 2014) scores >20, indicating potential deleterious effects on protein structure/function. Two missense alleles (S159A and A160P) are located within the *LOXL1* intrinsic disordered region (IDR) with high propensity to protein aggregation (**Figure 1**). All four missense variants were observed more often in controls compared to cases (**Table 1**) for both the XFS/XFG (combined P= 7.6E-4) and NEIGHBOR control datasets (combined P= 2.2E-3). Using data from an international XFS meta-analysis (Aung 2017), we assessed the allelic distributions for these variants in other populations. While these rare variants are not universally present, we were able to observe consistent protective effects in an Icelandic case-control study for A160P and S427F and in a South African cohort for S159A (**Table 2**). G120S was not associated (P> 0.05) with XFS in any other population (1 of 458 cases from Italy and 1 of 2827 cases from Japan and not in any controls from either population (267 and 3013, respectively).

**Table 1.**
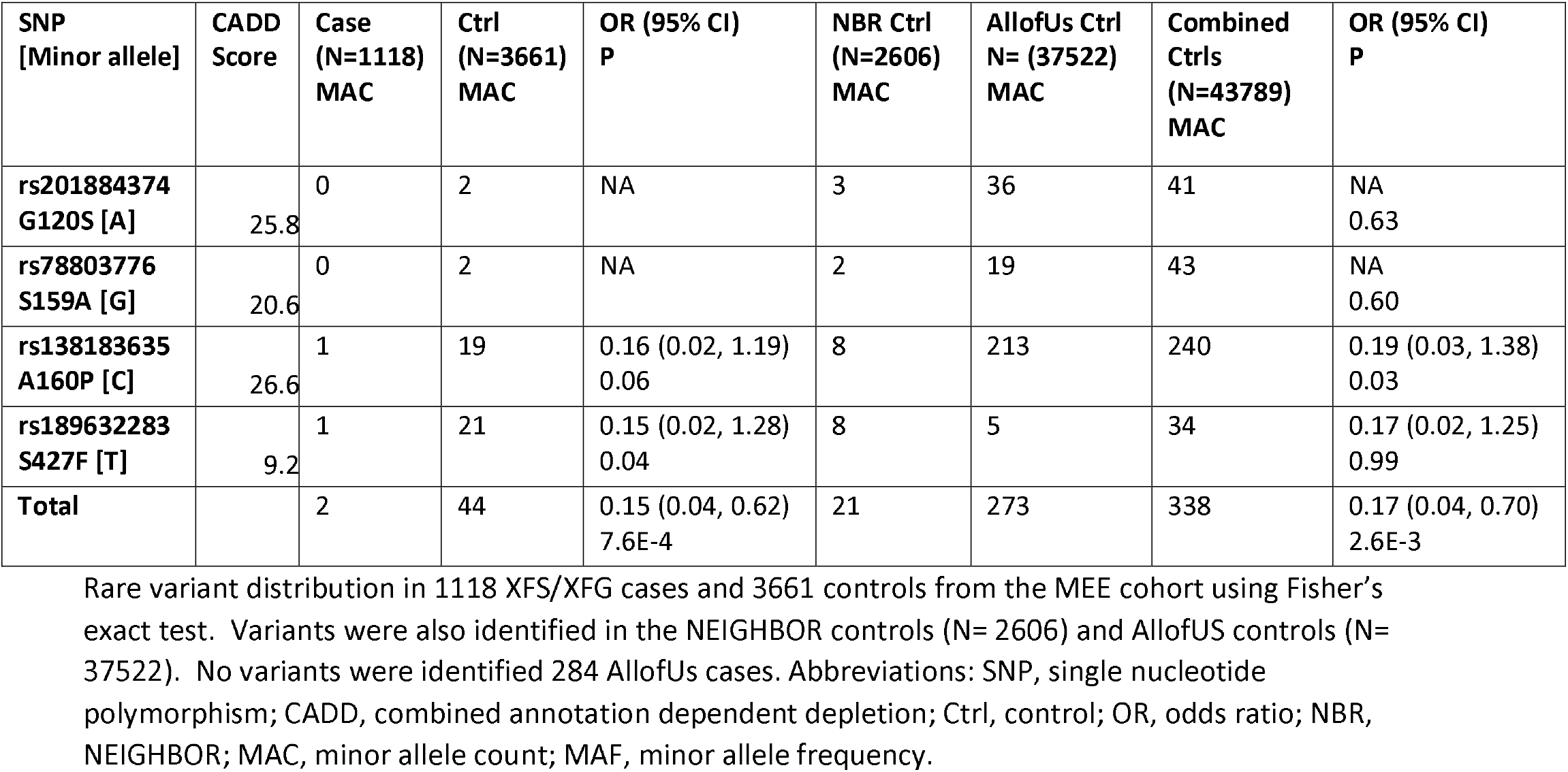
Distribution of rare *LOXL1* coding variants in XFS/XFG cases and controls in the MEE cohort.

**Table 2.** Rare *LOXL1* coding variant association in replication cohorts.

|  | SNP [Minor allele] |  |  |  |
| --- | --- | --- | --- | --- |
|  | rs201884374 G120S [A] | rs78803776 S159A [G] | rs138183635 A160P [C] | rs189632283 S427F [T] |
| Population | OR (P value) | OR (P value) | OR (P value) | OR (P value) |
| **South Africa**<br>90 cases<br>250 controls | NP | 0.21 (0.0004) | NP | NP |
| **Iceland**<br>195 cases<br>14,474 controls | NP | NP | 0.17 (0.0014) | 0.16 (0.026) |

rs78803776[G] was only present in the South African sample, while rs138183635[C] and rs189632283[T] were only present in the Icelandic sample. Abbreviations: OR, Odds ratio; NP, not present.

In AoU controls, 47 variants were found in 456 individuals (**Supplementary Table 2**). These are mainly missense alleles and were found only in controls and not in any of the 284 cases (P= 0.045). Three of the four *LOXL1* missense alleles identified in the MEE cohort were among the 47 variants (456 controls) identified in AllofUs controls: A160P (213 controls), S159A (19 controls) and G120S (36 controls) (**Supplementary Table 2**).

While XFS/XFG cases and controls could not be identified in the UK biobank, we did assess contributions of *LOXL1* rare coding variants in the UK Biobank glaucoma groups (‘Glaucoma H40’, ‘Glaucoma Custom’, ‘Glaucoma’, and ‘Glaucoma surgery’), (**Supplementary Table 3**) using the publicly available website Genebass (Karczewski, 2022). We examined each glaucoma group for aggregated variant effects (SKAT-O and Gene Burden) in 3 variant groups: loss of function alleles, missense alleles and synonymous alleles. For each phenotype group, nominal evidence of association was only observed for the missense alleles and in each case the effect was protective (**Supplementary Table 3**). Five alleles showed nominal single variant effects (**Supplementary Table 4**), including A160P with protective effects (P=5.29e-3 β=−3.29e-1).

### LOXL1 rare haplotypes are more common in controls than in cases

To further explore the protective effects of the rare missense alleles, in the MEE cohort, we investigated the distribution of haplotypes defined by the 4 missense alleles as well as the two common *LOXL1* variants, G153D and R141L. All haplotypes that include a rare variant allele or the minor allele for G153D and R141L are found more often in controls compared to cases (OR = 0.33, P=1.7E-173) (**Table 3**). In addition, only the haplotype that includes the common allele for all 6 missense variants was associated with increased disease risk (OR = 4.76, P= 1.7E-173).

**Table 3.** *LOXL1* haplotypes in the MEE cohort with common and rare coding variants and XFS/XFG association.

| Hap # | G120S | R141L | G153D | S159A | A160P | S427F | Cases<br>(N=2236) | Controls<br>(N=7322) | P | OR (95% CI) |
| --- | --- | --- | --- | --- | --- | --- | --- | --- | --- | --- |
|  |  |  |  |  |  |  | N | N |  |  |
| 1 | A | T | G | T | G | C | 0 | 2 | NA | NA |
| 2 | G | G | A | T | C | C | 1 | 19 | 0.06 | 0.16 (0.02, 1.19) |
| 3 | G | G | A | T | G | C | 34 | 1219 | 7.90E-106 | 0.08 (0.06, 0.11) |
| 4 | G | G | G | G | G | C | 0 | 2 | NA | NA |
| 5 | G | G | G | T | G | C | 1869 | 3785 | 1.7E-173 | 4.76 (4.20, 5.36) |
| 6 | G | T | G | T | G | C | 331 | 2274 | 3.74E-56 | 0.39 (0.34, 0.44) |
| 7 | G | T | G | T | G | T | 1 | 21 | 0.04 | 0.15 (0.02, 1.28) |
| All<br>minor |  |  |  |  |  |  | 367 | 3537 | 1.7E-173 | 0.21 (0.19, 0.24) |

The minor (rare) allele at each variant is highlighted in yellow. Only haplotype #5 (highlighted in orange) that includes all the major (common) alleles is associated with disease risk. Abbreviations: NA, not applicable; OR, odds ratio.

We also investigated haplotypes in the AoU data where only the haplotype homozygous for common ‘G’ alleles at G153D and R141L, without the addition of any rare alleles, was associated with disease risk (OR 3.22, P=6.2E-43) (Haplotype 1, **Table 5**).

### LOXL1 heterozygous diplotypes are more common in controls than in cases

As rare alleles are more likely to be present as heterozygous genotypes, in the MEE cohort, we examined the distribution of the 6-variant diplotypes (derived from the haplotypes in Table 3) among XFS/XFG cases and controls. Similar to the haplotype distribution, the homozygous diplotype comprised of the common alleles (Diplotype A, **Table 4**) is associated with increased disease risk (OR= 6.78, P=4.7E-157). All diplotypes that include heterozygous genotypes are protective with the exception of diplotype F, which was associated with an OR of 1.64 (**Table 4**) but was also very rare, and the distribution between cases and controls was not statistically significant (P= 0.55). Overall, heterozygous diplotypes are significantly protective (OR= 0.45, P=4.2E-89) (**Table 4**). We also observed similar protective effects for heterozygous diplotypes in the AllofUs data (**Table 6**) and an overall significant protective effect for heterozygous diplotypes (OR= 0.26, P=7.62E-28) (**Table 7**).

**Table 4.** *LOXL1* diplotypes and XFS/XFG association in the MEE cohort.

| Diplotype | G120S | R141L | G153D | S159A | A160P | S427F | Cases | Freq. | Controls | Freq. | P | OR (95% CI) |
| --- | --- | --- | --- | --- | --- | --- | --- | --- | --- | --- | --- | --- |
| A (H5/H5) | G | G | G | T | G | C | 786 | 0.703 | 947 | 0.259 | 4.7E-157 | 6.78 (5.87,7.89) |
|  | G | G | G | T | G | C |  |  |  |  |  |  |
| B (H6/H6) | G | T | G | T | G | C | 18 | 0.016 | 448 | 0.122 | 6.4E-34 | 0.13 (0.08,0.20) |
|  | G | T | G | T | G | C |  |  |  |  |  |  |
| Sum Homozygous |  |  |  |  |  |  | 804 | 0.719 | 1395 | 0.38 | 4.2E-89 | 4.16 (4.76,3.89) |
| C (H5/H6) | G | G | G | T | G | C | 290 | 0.259 | 1220 | 0.333 | 3.0E-06 | 0.7 (0.80,0.60) |
|  | G | T | G | T | G | C |  |  |  |  |  |  |
| D (H5/H3) | G | G | G | T | G | C | 15 | 0.013 | 659 | 0.18 | 1.8E-68 | 0.06 (0.10,0.04) |
|  | G | G | A | T | G | C |  |  |  |  |  |  |
| E (H7/H6) | G | T | G | T | G | T | 1 | 8.9E-4 | 7 | 1.9E-3 | 0.69 | 0.47 (3.78,0.48) |
|  | G | T | G | T | G | C |  |  |  |  |  |  |
| F (H2/H3) | G | G | A | T | C | C | 1 | 8.9E-4 | 2 | 5.5E-4 | 0.55 | 1.64 (0.15,1.64) |
|  | G | G | A | T | G | C |  |  |  |  |  |  |
| Sum one Heterozygous |  |  |  |  |  |  | 307 | 0.27 | 1888 | 0.516 | 5.7E-47 | 0.36 (0.41,0.31) |
| G (H3/H6) | G | G | A | T | G | C | 7 | 6.3E-3 | 343 | 0.093 | 4.7E-32 | 0.06 (0.13,0.03) |
|  | G | T | G | T | G | C |  |  |  |  |  |  |
| H (H5/H2) | G | G | G | T | G | C | 0 | 0 | 6 | 0.002 | 0.34 | NA |
|  | G | G | A | T | C | C |  |  |  |  |  |  |
| I<br>(H7/H5) | G | **T** | G | T | G | **T** | 0 | 0 | 13 | 0.004 | 0.048 | NA |
|  | G | **G** | G | T | G | **C** |  |  |  |  |  |  |
| J<br>(H6/H4) | G | **T** | G | **T** | G | C | 0 | 0 | 1 | 2.7E-4 | NA | NA |
|  | G | **G** | G | **G** | G | C |  |  |  |  |  |  |
| K<br>(H3/H4) | G | G | **A** | **T** | G | C | 0 | 0 | 1 | 2.7E-4 | NA | NA |
|  | G | G | **G** | **G** | G | C |  |  |  |  |  |  |
| L<br>(H1/H5) | **A** | **T** | G | T | G | C | 0 | 0 | 1 | 2.7E-4 | NA | NA |
|  | **G** | **G** | G | T | G | C |  |  |  |  |  |  |
| **Sum 2 heterozygous** |  |  |  |  |  |  | **7** | **0.0063** | **365** | **0.0997** | **1.2E-34** | **0.06 (0.13,0.** |
| M<br>(H2/H6) | G | **G** | **A** | T | **C** | C | 0 | 0 | 11 | 0.003 | 0.078 | NA |
|  | G | **T** | **G** | T | **G** | C |  |  |  |  |  |  |
| N<br>(H3/H7) | G | **G** | **A** | T | G | **C** | 0 | 0 | 2 | 5.5E-4 | NA | NA |
|  | G | **T** | **G** | T | G | **T** |  |  |  |  |  |  |
| **Sum 3 heterozygous** |  |  |  |  |  |  | **0** | **0** | **13** | **0.0036** | **0.048** | **NA** |
| **Sum 2 + 3 heterozygous** |  |  |  |  |  |  | **7** | **6.3E-3** | **378** | **0.10** | **1.0E-39** | **0.05 (0.10,0.** |
| **Sum all heterozygous** |  |  |  |  |  |  | **314** | **0.28** | **2266** | **0.62** | **4.2E-89** | **0.45 (0.52,0.** |

Heterozygous genotypes are shown in bold type. Only the homozygous diplotype A (H5/H5) is significantly associated with risk. Overall 72% of cases have a homozygous diplotype compared to 38% of controls. The diplotypes that include two heterozygous genotypes are found in 7 cases and 342 controls while the diplotypes with three heterozygous genotypes are only found in controls.

**Table 5.** *LOXL1* haplotypes in the AllofUs cohort with common and rare coding variants and XFS/XFG association.

| Haplotype Number | Haplotype | Frequency Cases (n=568) | Frequency Controls (n=74998) | OR | 95% CI | P |
| --- | --- | --- | --- | --- | --- | --- |
| 1 | GG | 447 (78.7%) | 38456 (51.3%) | 3.22 | 2.61-3.92 | 6.2E-43 |
| 2 | GG(+) | 0 (0%) | 139 (0.2%) | -- | -- | 0.41 |
| 3 | GG(++) | 0 (0%) | 2 (0.003%) | -- | -- | 0.07 |
| 4 | GA | 19 (3.3%) | 12039 (16.1%) | 0.14 | 0.09-0.22 | 5.1E-22 |
| 5 | GA(+) | 0 (0%) | 255 (0.3%) | -- | -- | 0.21 |
| 6 | TG | 102 (18.0%) | 24006 (32.0%) | 0.37 | 0.29-0.45 | 7.6E-14 |
| 7 | TG(+) | 0 (0%) | 101 (0.1%) | -- | -- | 0.54 |
| 8 | TA | 0 (0%) | 0 (0%) |  |  |  |
| 9 | TA(+) | 0 (0%) | 0 (0%) |  |  |  |
| All minor* |  | 121 (21.3%) | 36542 (48.7%) | 0.28 | 0.23-0.34 | 4.41E-41 |

\*Minor alleles include the rare alleles (MAF <1%) and the alternate alleles for the two common variants ([D] at G153D and [L] at R141L).

**Table 6.**
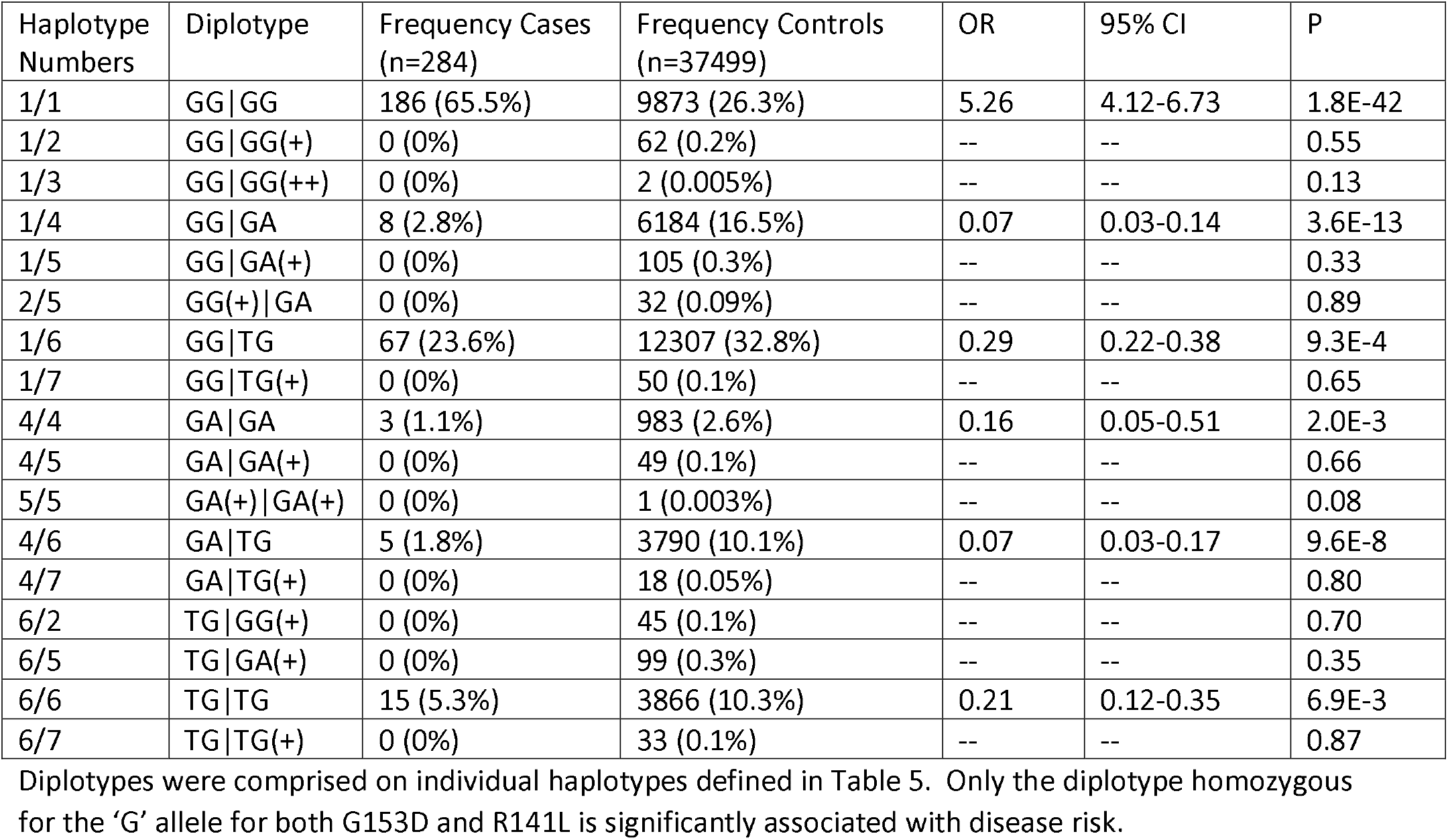
*LOXL1* diplotypes and XFS/XFG association in the AllofUs cohort.

| Haplotype Numbers | Diplotype | Frequency Cases (n=284) | Frequency Controls (n=37499) | OR | 95% CI | P |
| --- | --- | --- | --- | --- | --- | --- |
| 1/1 | GG \| GG | 186 (65.5%) | 9873 (26.3%) | 5.26 | 4.12-6.73 | 1.8E-42 |
| 1/2 | GG \| GG(+) | 0 (0%) | 62 (0.2%) | -- | -- | 0.55 |
| 1/3 | GG \| GG(++) | 0 (0%) | 2 (0.005%) | -- | -- | 0.13 |
| 1/4 | GG \| GA | 8 (2.8%) | 6184 (16.5%) | 0.07 | 0.03-0.14 | 3.6E-13 |
| 1/5 | GG \| GA(+) | 0 (0%) | 105 (0.3%) | -- | -- | 0.33 |
| 2/5 | GG(+) \| GA | 0 (0%) | 32 (0.09%) | -- | -- | 0.89 |
| 1/6 | GG \| TG | 67 (23.6%) | 12307 (32.8%) | 0.29 | 0.22-0.38 | 9.3E-4 |
| 1/7 | GG \| TG(+) | 0 (0%) | 50 (0.1%) | -- | -- | 0.65 |
| 4/4 | GA \| GA | 3 (1.1%) | 983 (2.6%) | 0.16 | 0.05-0.51 | 2.0E-3 |
| 4/5 | GA \| GA(+) | 0 (0%) | 49 (0.1%) | -- | -- | 0.66 |
| 5/5 | GA(+) \| GA(+) | 0 (0%) | 1 (0.003%) | -- | -- | 0.08 |
| 4/6 | GA \| TG | 5 (1.8%) | 3790 (10.1%) | 0.07 | 0.03-0.17 | 9.6E-8 |
| 4/7 | GA \| TG(+) | 0 (0%) | 18 (0.05%) | -- | -- | 0.80 |
| 6/2 | TG \| GG(+) | 0 (0%) | 45 (0.1%) | -- | -- | 0.70 |
| 6/5 | TG \| GA(+) | 0 (0%) | 99 (0.3%) | -- | -- | 0.35 |
| 6/6 | TG \| TG | 15 (5.3%) | 3866 (10.3%) | 0.21 | 0.12-0.35 | 6.9E-3 |
| 6/7 | TG \| TG(+) | 0 (0%) | 33 (0.1%) | -- | -- | 0.87 |

**Table 7.** Summary of homozygous and heterozygous diplotypes among cases and controls in AllofUs.

| Number of haplotypes per individual with homozygous or heterozygous diplotypes | Frequency Cases (n=284) | Frequency Controls (n=37499) | OR | 95% CI | P |
| --- | --- | --- | --- | --- | --- |
| All homozygous | 204 (72%) | 14,722 (39%) | 3.95 | 3.04- 5.11 | 3.0E-28 |
| 1 heterozygous | 75 (26%) | 18,553 (49%) | 0.37 | 0.28-0.48 | 2.4E-16 |
| 2 heterozygous | 5 (0.18%) | 4125 (11%) | 0.15 | 0.06- 0.35 | 4.6E-9 |
| 1+2 heterozygous | 80 (28%) | 22,678 (60%) | 0.26 | 0.18-0.33 | 7.6E-28 |

Summary of the diplotypes shown in Table 7 according to number of heterozygous genotypes.

Two diplotypes comprised of only homozygous genotypes had higher frequency in controls: the GA|GA diplotype in AllofUs (**Table 6**) and the TG|TG diplotype in Allof Us and corresponding diplotype B in the MEE cohort (**Table 4 and Table 6**). Both of these diplotypes include the homozygous minor allele for one of the two common variants.

### Distribution of heterozygous variants with protective effects in LOXL1 disordered domains

We used the disordered domain predictor PONDR (Peng, 2005) to define the LOXL1 disordered regions (**Figure 1**). The variants with protective effects were preferentially located in the LOXL1 disordered region extending from amino acid residue 138 to 166 (**Table 8**) (P= 5.81E-45).

**Table 8.** Enrichment of rare variants in controls in predicted disordered segments.

| Predicted Disorder Segment<br>(Amino acid residue) | Number of rare alleles<br>N=456 | P-value |
| --- | --- | --- |
| 1-8 | 3 | 0.36 |
| 56-109 | 40 | 0.82 |
| 138-166 | 241 | 5.8E-45 |
| 212-226 | 0 | -- |
| 251-276 | 0 | -- |
| 286-380 | 56 | 0.11 |

### Excess homozygosity and decreased heterozygosity for common variants in XFS/XFG cases

Considering the consistent protective effects for heterozygous variants, we hypothesized that in case/control cohorts, an excess of homozygosity for the common G153D and R141L variants could be observed in cases (with a decrease in heterozygosity) and an excess of heterozygosity could be evident in controls. To investigate this hypothesis, we used genotype data from 40 independent previously published studies for 3 genetic ancestry groups (European, Asian, and African) summarized in Li et al (Li, 2021), and calculated the expected and observed distribution of genotype groups using Hardy Weinberg equilibrium and population-specific allele frequencies. We found that in all ancestral groups there was an excess of homozygosity and a decrease in heterozygosity for G153D genotypes in cases (Supplemental Table 5A). A similar trend was observed for the G153D in AllofUs (Europeans) and in the MEE cohort (Supplemental Table 5B). The combined data for G153D in cases (N=5748) showed a significant excess of homozygosity for both the major and minor alleles and a significant decrease in heterozygosity (Table 9). For R141L, similar deviations were observed for the Korean and Japanese cases N=1039 (Table 10) but not for any other genetic ancestry or the AoU or MEE cohorts (Supplementary table 6). In controls, only the data from the MEE cohort showed an excess of heterozygote genotypes for G153D (Supplementary Table 5B).

**Table 9.** rs3825942 (G153D) genotype distribution in cases (N=5748) from the combined datasets.

|  |  |  |  |  |
| --- | --- | --- | --- | --- |
| **All genotypes** | GG | GA | AA |  |
| Observed | 5383 | 311 | 49 |  |
| Expected | 5297 | 397 | 7.5 | P=1.0e-6 |
| **All Homozygous** | GG+AA |  |  |  |
| Observed | 5514 |  |  |  |
| Expected | 5414 | P=1.9E-4 |  |  |
| **Homozygous minor allele** | AA+GG* |  |  |  |
| Observed | 56 |  |  |  |
| Expected | 10 | P=6.3E-9 |  |  |
| **Heterozygous** | GA |  |  |  |
| Observed | 311 |  |  |  |
| Expected | 392 | 1.8E-3 |  |  |

**Table 10.**
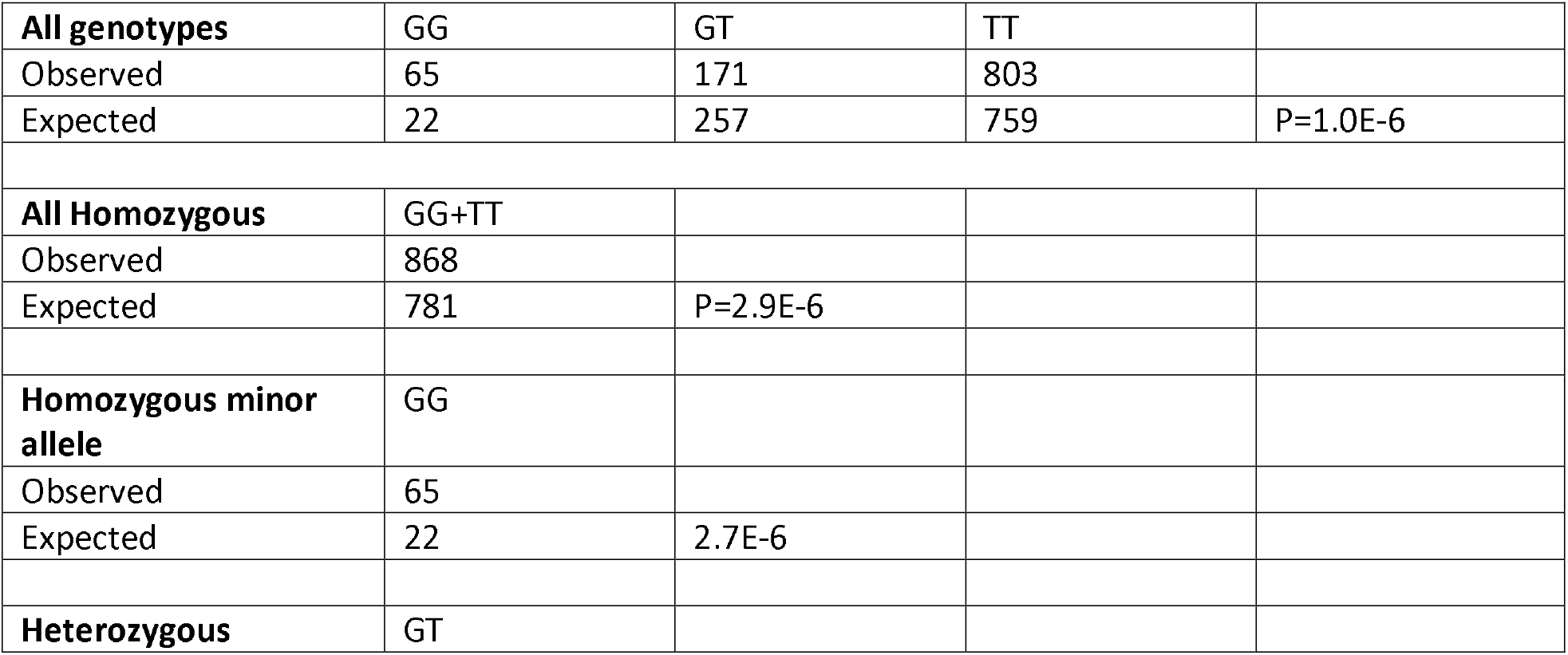
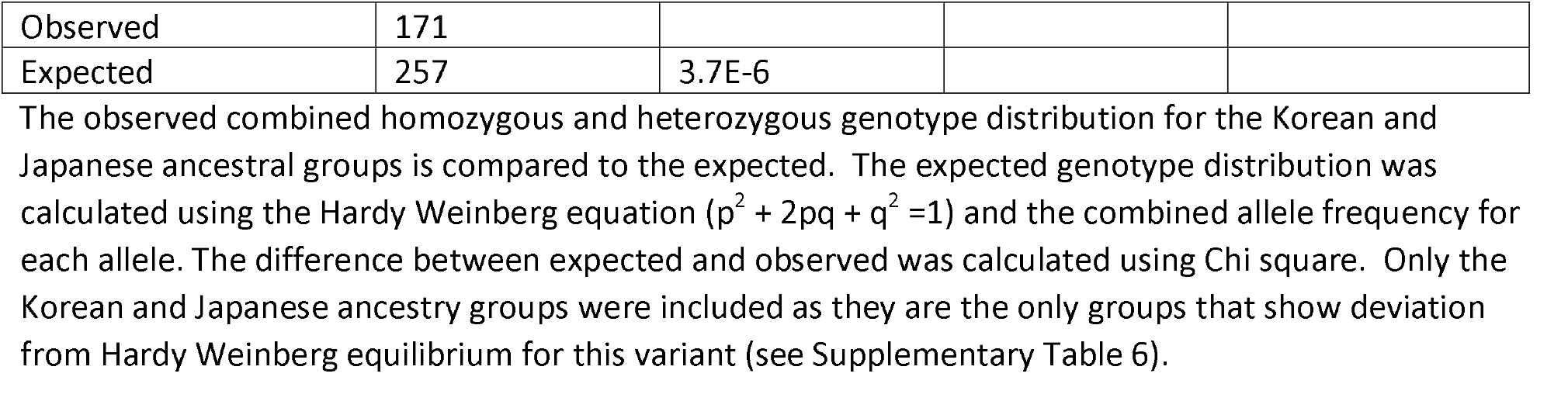
rs1048661 (R141L) genotype distribution in Japanese and Korean cases (N=1039)

|  |  |  |  |  |
| --- | --- | --- | --- | --- |
| **All genotypes** | GG | GT | TT |  |
| Observed | 65 | 171 | 803 |  |
| Expected | 22 | 257 | 759 | P=1.0E-6 |
| **All Homozygous** | GG+TT |  |  |  |
| Observed | 868 |  |  |  |
| Expected | 781 | P=2.9E-6 |  |  |
| **Homozygous minor allele** | GG |  |  |  |
| Observed | 65 |  |  |  |
| Expected | 22 | 2.7E-6 |  |  |
| **Heterozygous** | GT |  |  |  |

|  |  |  |
| --- | --- | --- |
| Observed | 171 |  |
| Expected | 257 | 3.7E-6 |

## DISCUSSION

Common *LOXL1* coding variants are major XFS/XFG genetic risk factors, but despite the strong association with disease risk, the underlying pathogenic mechanism(s) are not known. The discovery of a protective rare *LOXL1* variant in a Japanese case-control study provided the first insight that *LOXL1* protein-altering variants could reduce disease risk. In this study, we have expanded this initial observation to include other rare *LOXL1* missense alleles that are all more commonly found in controls compared to cases. In particular, A160P shows protective effects in the MEE cohort, an Icelandic dataset, AoU, and the UK Biobank. We also show that haplotypes and diplotypes that include these rare coding variants are more common in controls.

In case/control studies, deviation from Hardy Weinberg distribution in cases for the common variants G153D and R141L suggests that overall, heterozygosity is protective and homozygosity is associated with increasing disease risk. We did not observe a consistent significant deviation from expected patterns in controls, which could be due to several factors, including undiagnosed cases in controls. Interestingly, the only control group with excess heterozygosity was the MEE cohort, where controls had eye exams and did not report having a physician-diagnosis of glaucoma.

The risk effect for homozygous variants is supported by the observation that only the diplotype that includes common homozygous variants (diplotype A in the MEE cohort and diplotype GG|GG in AoU) is associated with increased disease risk. However, the homozygous diplotypes that include homozygosity for the minor alleles G153D [D] and R141L [L], (diplotype B in MEE and diplotypes GA|GA and TG|TG in AllofUs), while rare, are found more often in controls, potentially suggesting that homozygosity alone is not sufficient for disease development. It is not clear what factors could lead to the higher frequency of these diplotypes in controls; however, it is intriguing that the homozygous ‘minor’ allele is involved in both diplotypes demonstrating this effect. It is possible that other, as yet undetected rare variants, are present on these diplotypes that are underlying the protective effects, especially considering that these diplotypes are not common in any population. Interestingly, both homozygous ‘D’ and homozygous ‘L’ are more common in combined cases than expected for G153D and in the Korean/Japanese datasets for R141L (**Tables 9, 10**).

Rare variants with protective effects are primarily located in a LOXL1 disordered region that includes the ‘common’ variants G153D and R141L, and includes a region that has been shown to reduce aggregation when deleted from the protein (Bernstein 2019). This observation, coupled with the protective effects of heterozygosity, suggests that protein variation within this region could reduce protein aggregation. IDRs can contribute to pathogenic aggregation for several neurodegenerative diseases including Alzheimer’s, ALS, Parkinson’s, Huntington’s, and prion-related diseases, among others (Eftekharzadeh 2016, Flores, 2025). Understanding the nature of aggregation induced by IDRs is an area of intense investigation (Flores, 2025). Protein variants may increase or decrease aggregation (Candelise 2021), with specific examples including a missense allele protective for Alzheimer’s disease shown to reduce amyloid beta aggregation (Das, 2017) and TDP-43 variants that influence the rate of protein aggregation related to ALS (Jiang, 2016). The strong homozygous and heterozygous effects observed in this study suggest that a matched amino acid sequence within the LOXL1 IDR supports aggregation (increases risk) while a mismatch (heterozygous) reduces aggregation and reduces risk.

In some populations, the risk allele at the common *LOXL1* variants (G153D and R141L) are reversed making it difficult to evoke a specific biological mechanism that explains disease risk. For example, a molecular interaction with the allele associated with risk or allele-specific gene expression influencing risk would have to be population dependent. While possible, it seems unlikely that a gene variant could underlie a very different function in one population compared to another. The results from this study, which showed a strong correlation between homozygosity and disease risk combined with overall protective effects observed for heterozygous diplotypes, supports a novel hypothesis that the observed risk allele reversal in some populations is simply the result of variation in minor allele frequency due to population effects and leading to variation in relative homozygosity and heterozygosity of risk variants. For example, R141 allele ‘L’ is the minor allele in most populations and is also associated with protective effects in most populations. However, in East Asians, where ‘L’ is the risk allele (Hayashi 2008), the ‘L’ allele is the more frequent allele and therefore is more likely to be homozygous and associated with disease risk. A similar argument can be made for the reversed risk observed for the G153D variant in South Africans (Williams 2010). This hypothesis also explains the protective effects of rare *LOXL1* protein-coding variants, as these are likely to be heterozygous due to their low minor allele frequencies. Further work will be required to confirm this hypothesis including testing heterozygous versus homozygous variants for protective functional effects.

There are several limitations of our study. First, the MEE analyses are based on exome array data, which does not comprehensively sample all *LOXL1* variants. The comprehensive sequence data fromAoU does help mitigate this concern; however, the number of cases in AoU is relatively small. Further work using larger numbers of cases with exome data will be necessary to further define the effects of rare coding variants on disease risk. Second, for the datasets available for this study, the number of controls is larger than the number of cases, which may provide more power to detect rare variants in controls. However, the decrease in heterozygosity for the common variants suggests that the higher frequency of heterozygous variants in controls is not entirely due to the larger sample size. Additionally, the significant enrichment of the AoU variants in the IDR argues against random distribution of rare variants in controls. Third, the differences in average ages for cases and controls may influence these results; however, the overall difference in age was not significant, and the distribution of diplotypes among case and control age groups was not significantly different (Supplementary Figure 1). Finally, because only one haplotype including a rare variant did not also include the protective allele for the common variants, we could not examine the effects of rare variants independent of the common risk alleles.

In summary, we identified rare protective *LOXL1* protein coding variants and showed that the distribution of the haplotypes and diplotypes that include these variants suggest an association with reduced risk of XFS/XFG. We also showed that only a homozygous diplotype comprised of all common alleles was significantly associated with disease risk and that excess homozygosity of the common risk variants G153D [G] and R141L [R] was observed in cases across ancestral groups. These results support a novel hypothesis that the reversal of *LOXL1* risk alleles observed in some populations is dependent on variation of minor allele frequencies among populations and that *LOXL1* allelic homozygosity is associated with increased disease risk, potentially due to effects on protein aggregation involving intrinsic disordered regions. Further work identifying additional protective variants and testing effects on protein aggregation will be necessary to confirm this hypothesis.

## Data Availability

All data produced in the present study are available upon reasonable request to the authors. Exome array data for this study has been deposited in dbGaP.

## ACKNOWLEDGEMENTS

Supported by NIH/NEI R01 EY020928, EY015473, and an unrestricted challenge grant from Research to Prevent Blindness (NYC to Icahn School of Medicine at Mount Sinai). Dr. Pasquale is supported by The Barry Family Center for Ophthalmic Artificial intelligence and The Glaucoma Foundation in New York City.

## Supplementary Tables

**Supplementary Table 1.** XFS/XFG case and control demographics.

|  | MEE cases | MEE controls | NEIGHBOR controls | AllofUs cases | AllofUs controls | P value |
| --- | --- | --- | --- | --- | --- | --- |
| N | 1118 | 3661 | 2606 | 284 | 37522 |  |
| % Female | 69 | 65 | 64 | 60 | 51 | 0.003 |
| Age (average) | 72.9 ± 9.6 | 54.0 ± 7.8 | 65.5 ± 13.5 | 80.5 ± 8.1 | 80.2 ± 4.6 | 0.28 |

**Supplementary Table 2.**
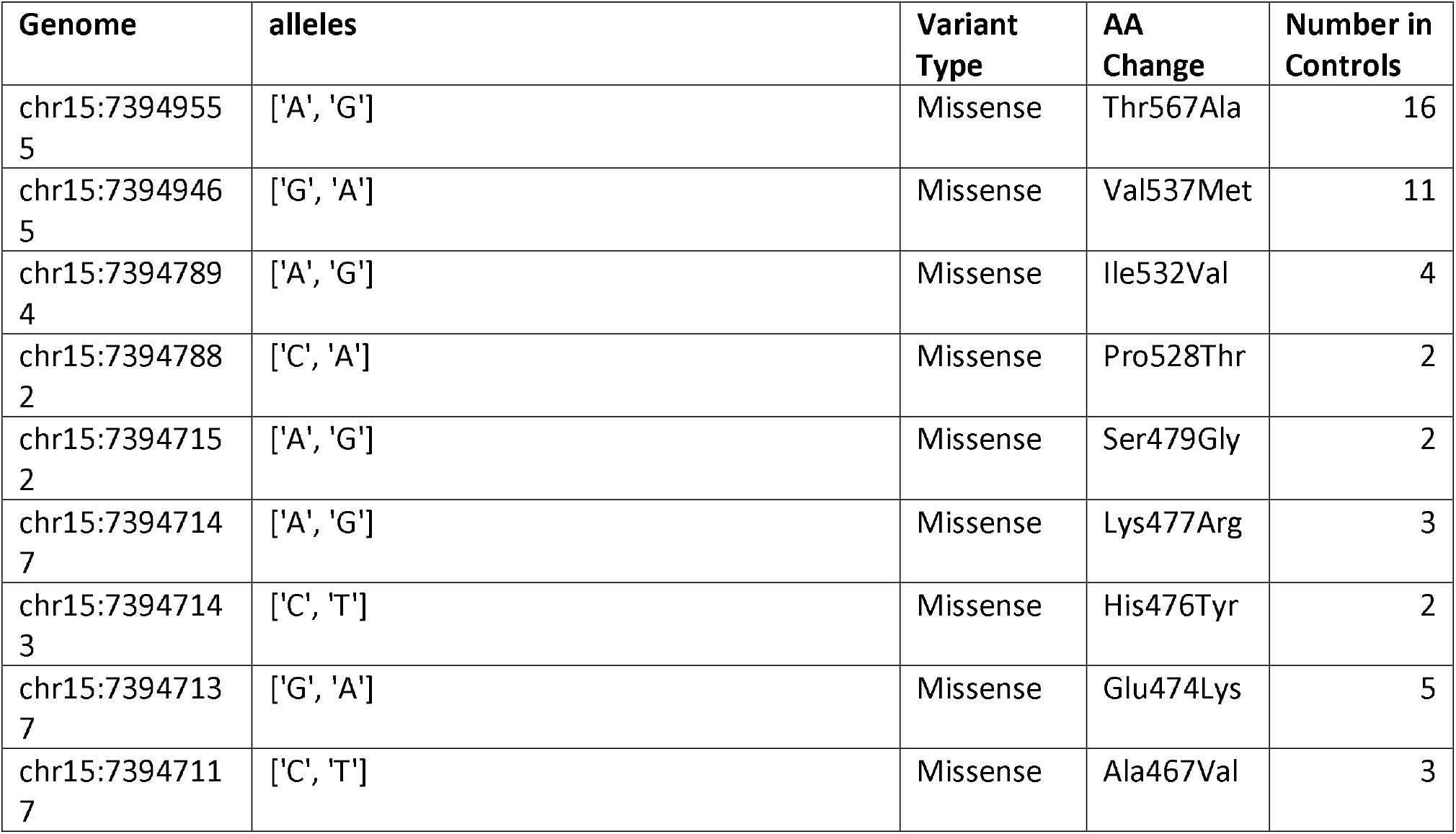
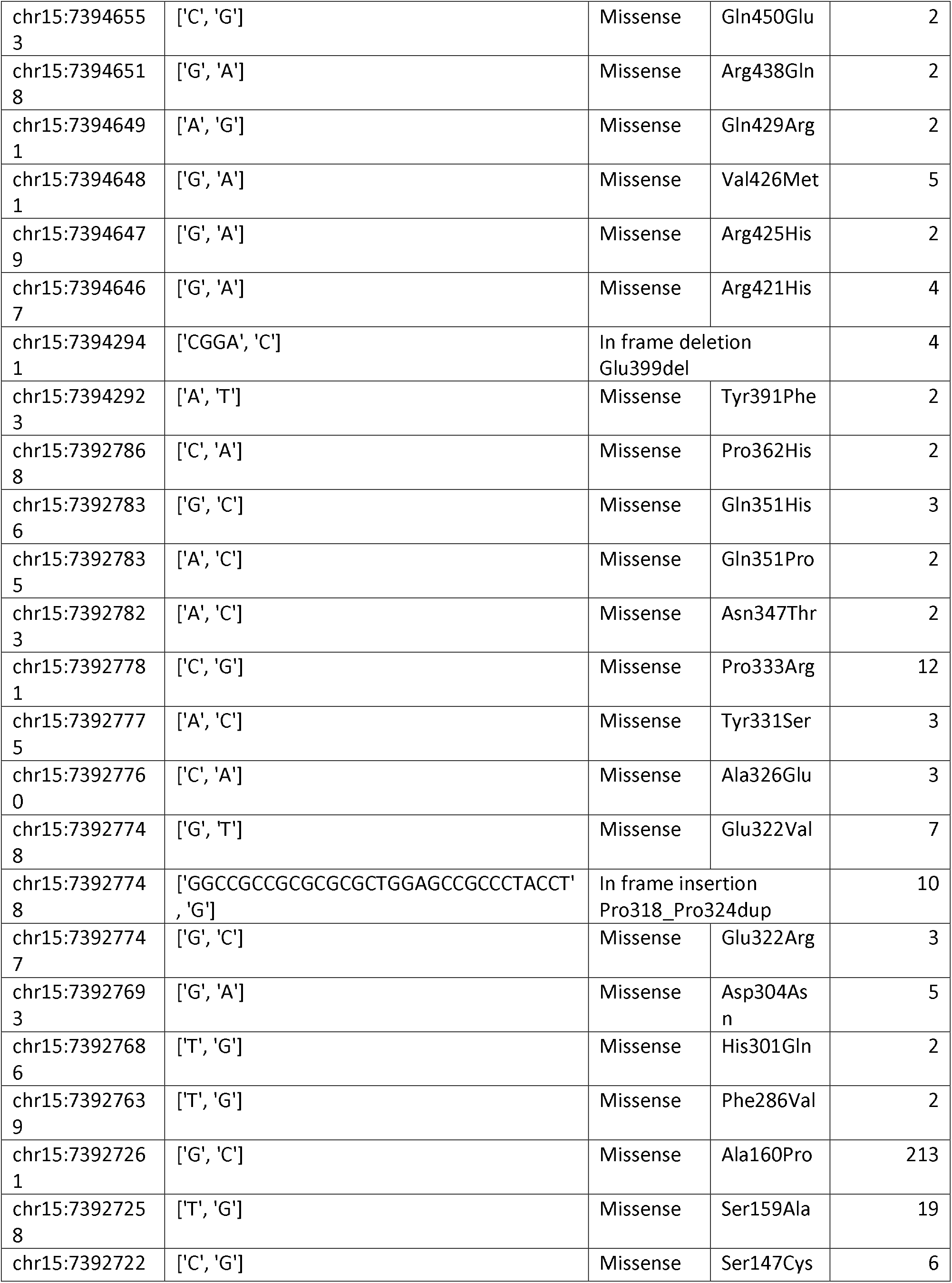
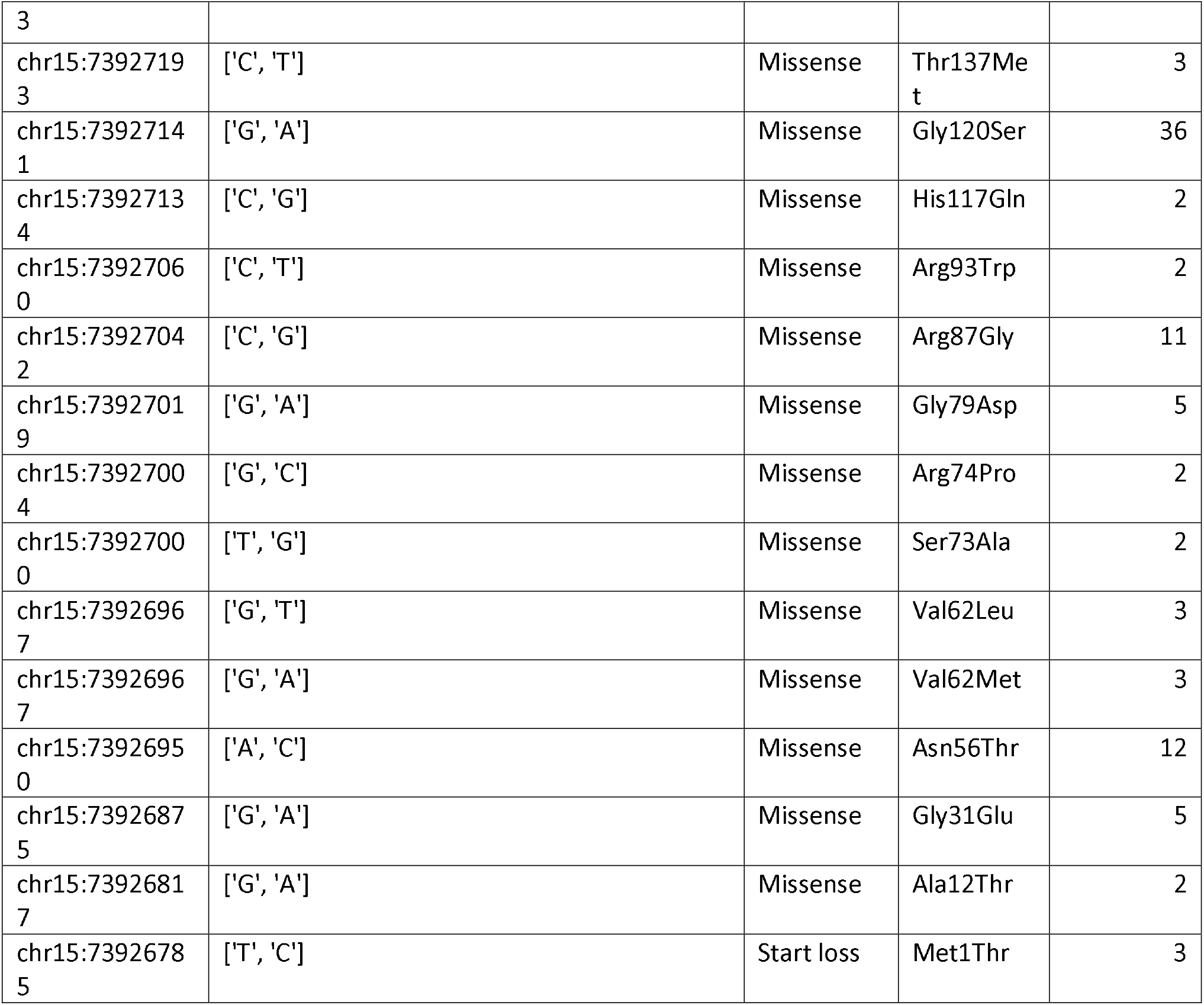
LOXL1 alleles identified in the *All of Us* cohort.

**Supplementary Table 3.** *LOXL1* exome variant association in UK Biobank.

| Dataset | SKAT-O |  |  | Burden |  |  |
| --- | --- | --- | --- | --- | --- | --- |
|  | LoF<br>N=30 | Missense<br>N=361 | Synonymous<br>N=189 | LoF<br>N=30 | Missense<br>N=361 | Synonymous<br>N=189 |
| Glaucoma H40<br>(11588 cases,<br>383253 controls) | P= 0.577<br>$\beta$= -2.3e-2 | P= 3.39e-3<br>$\beta$= -8.37e-3 | P= 0.374<br>$\beta$= 5.82e-3 | P= 0.587<br>$\beta$= -2.3e-2 | P= 1.82e-3<br>$\beta$= -8.7e-3 | P= 0.371<br>$\beta$= 5.82e-3 |
| Glaucoma custom<br>8912 cases,<br>385929, controls | P= 0.783<br>$\beta$= -1.69e-2 | P= 8.55e-3<br>$\beta$= -9.03e-3 | P= 0.632<br>$\beta$= 2.06e-3 | P= 0.565<br>$\beta$= -1.69e-2 | P= 4.33e-3<br>$\beta$= -9.03e-3 | P= 0.781<br>$\beta$= 2.06e-3 |
| Glaucoma<br>4063 cases,<br>390720 controls | P= 0.504<br>$\beta$= 8.353e-3 | P= 0.013<br>$\beta$= -1.24e-2 | P= 0.859<br>$\beta$= -1.62e-3 | P= 0.841<br>$\beta$= 8.35e-3 | P= 7.93e-3<br>$\beta$= -1.24e-2 | P= 0.879<br>$\beta$= 2.01e-3 |
| Glaucoma<br>Surgery<br>560 cases,<br>394223 controls | P= 1.73e-3<br>$\beta$= 0.178 | P= 0.127<br>$\beta$= -2.23e-2 | P= 0.654<br>$\beta$= 2.01e-3 | P= 0.0473<br>$\beta$= 1.78e-1 | P= 7.25e-2<br>$\beta$= -2.23e-2 | P= 0.941<br>$\beta$= -1.62e-3 |

Gene burden results (SKAT-O and Burden) in the UK Biobank were assessed using GeneBass (Karczewski, 2022) for all 4 glaucoma phenotype groups and three variant classification groups (Loss of function, LoF, Missense and Synonymous). N= the number of variants in each group. For all glaucoma groups and for both gene burden methods the missense variants are consistently protective.

**Supplementary Table 4.** Individual variant results for *LOXL1* variants in the UK Biobank.

| Variant | IDR | Glaucoma H40 | Case AC<br>N= 11588 | Control AC<br>N= 383253 |
| --- | --- | --- | --- | --- |
| Asn56Thr | Y (56-109) | P= 5.04e-2<br>$\beta$= -7.53e-1 | 2 | 234 |
| Ala160Pro | Y (138-166) | P=5.29e-3<br>$\beta$= -3.29e-1 | 48 | 2430 |
| Pro293Ser | Y (286-380) | P=5.1e-2<br>$\beta$= 4.3 | 1 | 2 |
| Cys401Tyr | N | P=4.74e-2<br>$\beta$= 4.13 | 1 | 2 |
| Leu544Ser | N | P=4.58e-2<br>$\beta$= 4.17 | 1 | 2 |

Variants with allele count (AC) of at least 2 in cases+controls, and p values for association of <0.05 are listed. Asn56Thr, Ala160Pro, Pro293Ser are all located within *LOXL1* intrinsic disordered regions (IDR).

**Supplementary Table 5.**
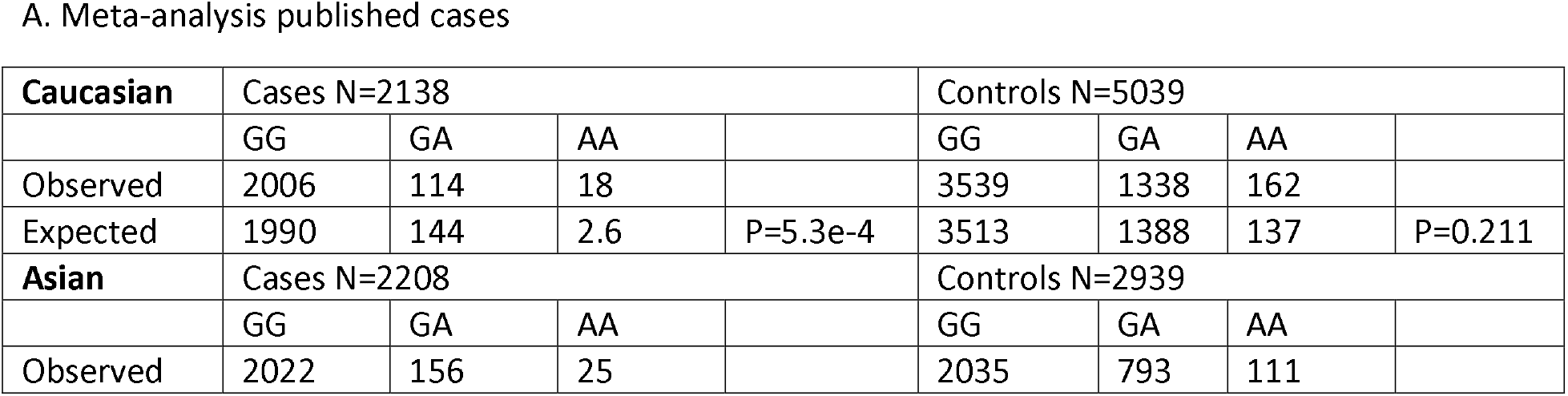
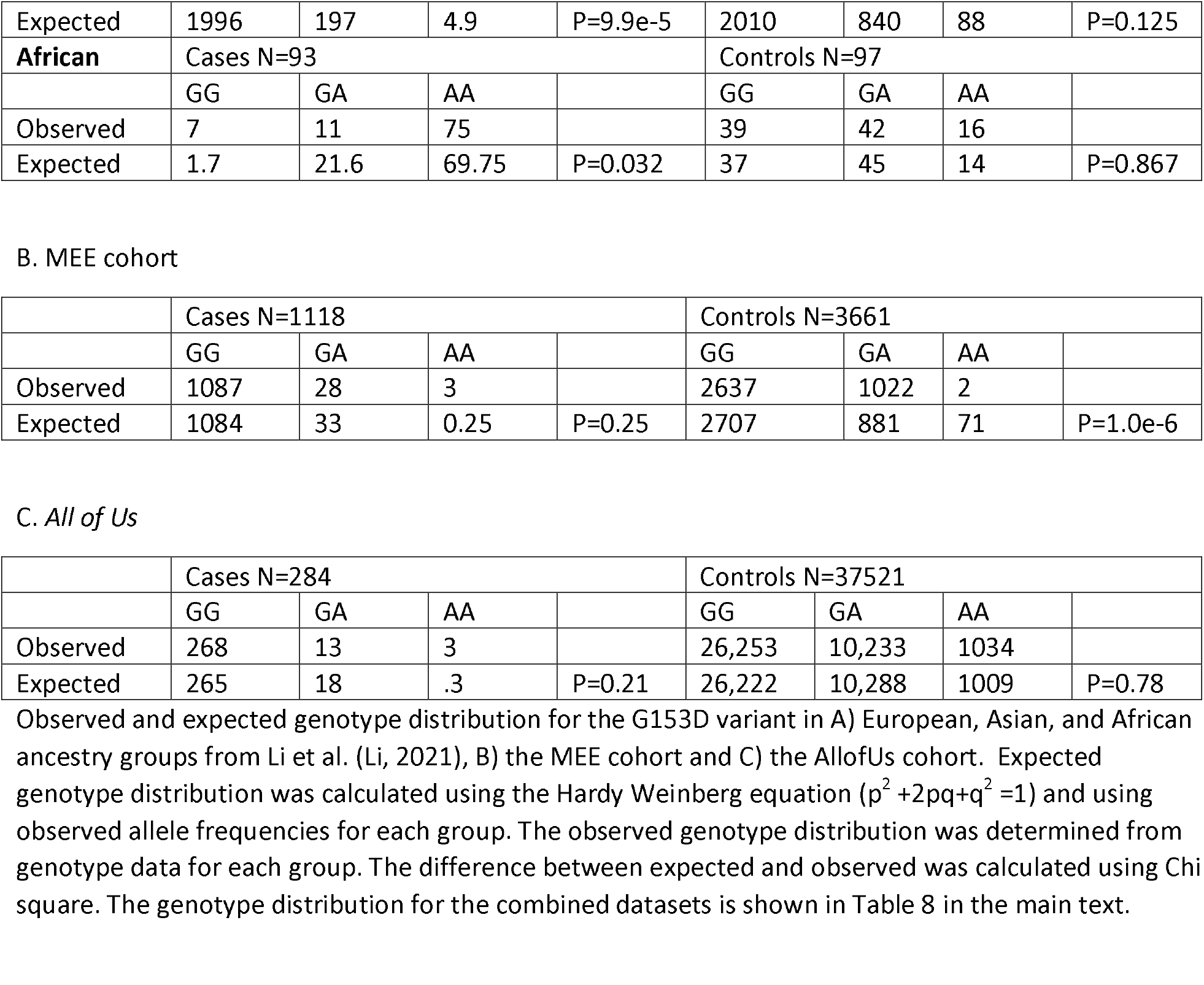
Observed and expected genotype distribution for G153D (rs3825942)

A. Meta-analysis published cases
| **Caucasian** | Cases N=2138 |  |  |  | Controls N=5039 |  |  |  |
| --- | --- | --- | --- | --- | --- | --- | --- | --- |
|  | GG | GA | AA |  | GG | GA | AA |  |
| Observed | 2006 | 114 | 18 |  | 3539 | 1338 | 162 |  |
| Expected | 1990 | 144 | 2.6 | P=5.3e-4 | 3513 | 1388 | 137 | P=0.211 |
| **Asian** | Cases N=2208 |  |  |  | Controls N=2939 |  |  |  |
|  | GG | GA | AA |  | GG | GA | AA |  |
| Observed | 2022 | 156 | 25 |  | 2035 | 793 | 111 |  |
| Expected | 1996 | 197 | 4.9 | P=9.9e-5 | 2010 | 840 | 88 | P=0.125 |
| **African** | Cases N=93 |  |  |  | Controls N=97 |  |  |  |
|  | GG | GA | AA |  | GG | GA | AA |  |
| Observed | 7 | 11 | 75 |  | 39 | 42 | 16 |  |
| Expected | 1.7 | 21.6 | 69.75 | P=0.032 | 37 | 45 | 14 | P=0.867 |
| B. MEE cohort |  |  |  |  |  |  |  |  |
|  | Cases N=1118 |  |  |  | Controls N=3661 |  |  |  |
|  | GG | GA | AA |  | GG | GA | AA |  |
| Observed | 1087 | 28 | 3 |  | 2637 | 1022 | 2 |  |
| Expected | 1084 | 33 | 0.25 | P=0.25 | 2707 | 881 | 71 | P=1.0e-6 |
| C. All of Us |  |  |  |  |  |  |  |  |
|  | Cases N=284 |  |  |  | Controls N=37521 |  |  |  |
|  | GG | GA | AA |  | GG | GA | AA |  |
| Observed | 268 | 13 | 3 |  | 26,253 | 10,233 | 1034 |  |
| Expected | 265 | 18 | .3 | P=0.21 | 26,222 | 10,288 | 1009 | P=0.78 |

**Supplementary Table 6.** Observed and expected genotype distribution for R141L (rs1048661)

A. Meta-analysis published cases
|  |  |  |  |  |  |  |  |  |
| --- | --- | --- | --- | --- | --- | --- | --- | --- |
| **Caucasian** | Cases N=2596 |  |  |  | Controls N=5039 |  |  |  |
|  | GG | GT | TT |  | GG | GT | TT |  |
| Observed | 1727 | 805 | 64 |  | 2530 | 2437 | 612 |  |
| Expected | 1745 | 766 | 84 | P=0.15 | 2504 | 2467 | 607 | P=0.33 |
| **Asian\*** | Cases N=1120 |  |  |  | Controls N=1474 |  |  |  |
|  | GG | GT | TT |  | GG | GT | TT |  |
| Observed | 738 | 324 | 58 |  | 635 | 647 | 192 |  |
| Expected | 723 | 352 | 43 | P=0.17 | 622 | 668 | 179 | P=0.63 |
| **Japanese and Korean** | Cases N=1039 |  |  |  | Controls N=1323 |  |  |  |
|  | GG | GT | TT |  | GG | GT | TT |  |
| Observed | 65 | 171 | 803 |  | 311 | 684 | 328 |  |
| Expected | 22 | 257 | 759 | P=1.0e-6 | 322 | 661 | 338 | P=0.69 |
| **African** | Cases N=93 |  |  |  | Controls N=97 |  |  |  |
|  | GG | GT | TT |  | GG | GT | TT |  |
| Observed | 92 | 1 | 0 |  | 70 | 24 | 3 |  |
| Expected | 78 | 2 | 0 | P=0.53 | 69 | 25 | 2 | P=0.89 |

B. MEE cohort
|  | Cases N=1118 |  |  |  | Controls N=3661 |  |  |  |
| --- | --- | --- | --- | --- | --- | --- | --- | --- |
|  | GG | GT | TT |  | GG | GT | TT |  |
| Observed | 802 | 297 | 19 |  | 1615 | 1591 | 455 |  |
| Expected | 807 | 285 | 25 | P=0.58 | 1585 | 1648 | 428 | P=0.34 |

C. All of Us
|  | Cases N=284 |  |  |  | Controls N=37521 |  |  |  |
| --- | --- | --- | --- | --- | --- | --- | --- | --- |
|  | GG | GT | TT |  | GG | GT | TT |  |
| Observed | 197 | 72 | 15 |  | 17306 | 16314 | 3901 |  |
| Expected | 190 | 84 | 9.2 | P=0.29 | 17350 | 16329 | 3842 | P=0.77 |

**Supplementary Figure 1.**
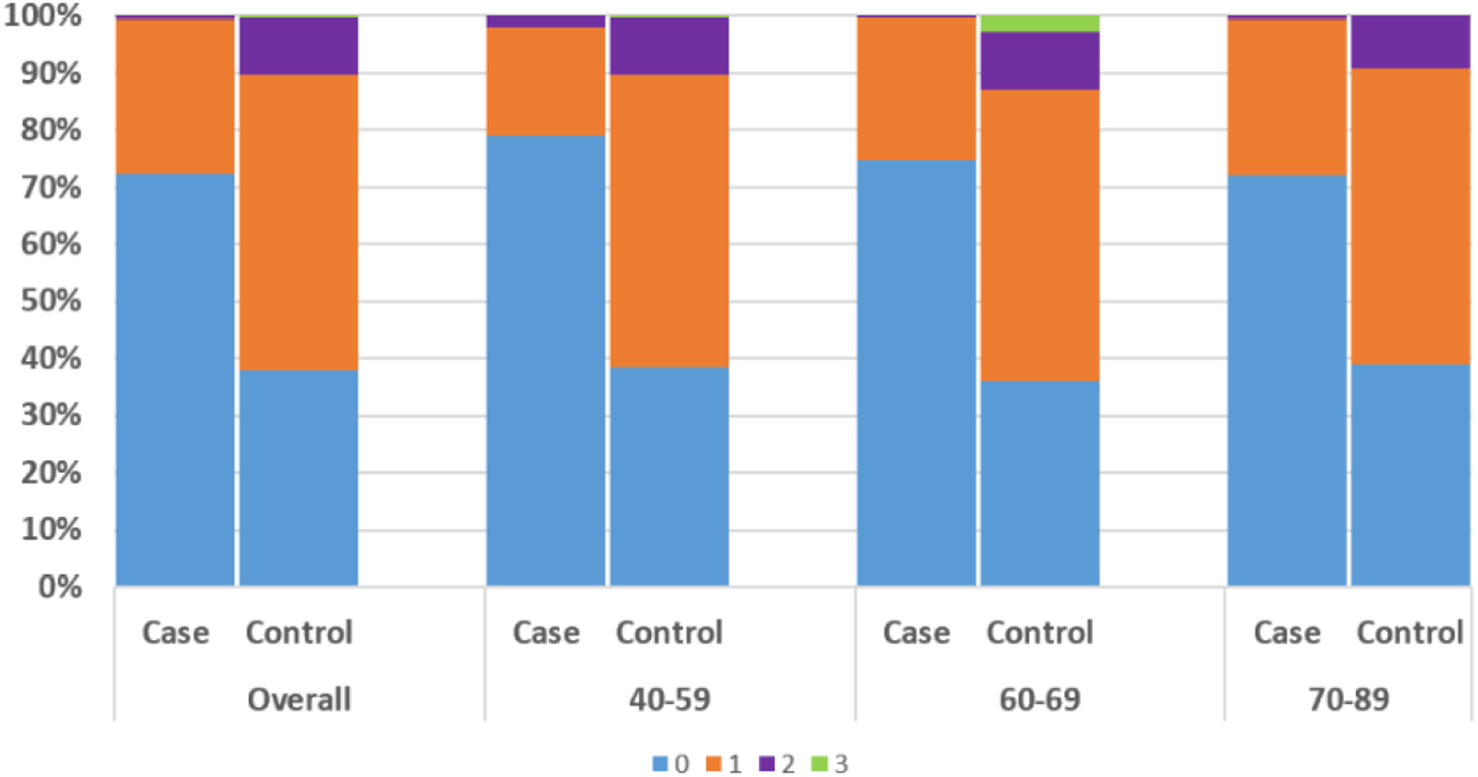
*LOXL1* diplotype distribution among cases and controls according to age. Total diplotypes with 0 heterozygous genotypes (ie homozygous) (blue), 1 heterozygous genotype (orange), 2 heterozygous genotypes (purple) and 3 heterozygous genotypes (green) were summed for cases and controls according to age groups: 40-59, 60-69, 70-89 and compared with the distribution among cases and controls overall. Similar distribution of diplotypes was observed for all age groups (P>0.05) with the exception of the rare diplotypes with 3 heterozygous variants which were found only in controls primarily in the 60-69 age range.

